# Longitudinal Analysis Over Decades Reveals the Development and Immune Implications of Type I Interferon Autoantibodies in an Aging Population

**DOI:** 10.1101/2024.02.27.24303363

**Authors:** Sonja Fernbach, Nina K. Mair, Irene A. Abela, Kevin Groen, Roger Kuratli, Marie Lork, Christian W. Thorball, Enos Bernasconi, Paraskevas Filippidis, Karoline Leuzinger, Julia Notter, Andri Rauch, Hans H. Hirsch, Michael Huber, Huldrych F. Günthard, Jacques Fellay, Roger D. Kouyos, Benjamin G. Hale, The Swiss HIV Cohort Study

**Affiliations:** Institute of Medical Virology, University of Zurich, Zurich, Switzerland; Department of Infectious Diseases and Hospital Epidemiology, University Hospital Zurich, University of Zurich, Zurich, Switzerland; Precision Medicine Unit, Lausanne University Hospital and University of Lausanne, Lausanne, Switzerland; Division of Infectious Diseases, Ente Ospedaliero Cantonale Lugano, University of Geneva and University of Southern Switzerland, Lugano, Switzerland; Infectious Diseases Service, Department of Medicine, Lausanne University Hospital and University of Lausanne, Lausanne, Switzerland; Clinical Virology, University Hospital Basel, Basel, Switzerland; Division of Infectious Diseases, Cantonal Hospital St. Gallen, St. Gallen, Switzerland; Department of Infectious Diseases, Inselspital, Bern University Hospital, University of Bern, Bern, Switzerland; Transplantation and Clinical Virology, Department of Biomedicine, University of Basel, Basel, Switzerland; School of Life Sciences, École Polytechnique Fédérale de Lausanne, Lausanne, Switzerland

## Abstract

Pre-existing autoantibodies (autoAbs) neutralizing type I interferons (IFN-Is: IFNα, IFNβ, IFNω) have recently been described as significant contributors to the severity of viral infectious diseases. Here, we explore the development and consequences of anti-IFN-I autoAbs at high-resolution using retrospective samples and data from 1876 well-treated individuals >65 years of age enrolled in the Swiss HIV Cohort Study, a nationwide, longitudinal cohort with up to 35 years of follow-up. Approximately 1.9% of individuals developed anti-IFN-I autoAbs, with a median onset age of ∼63 years (range 45-80). Once developed, anti-IFN-I autoAbs persisted for life, and generally increased in titer over years. Most individuals developed distinct neutralizing and non-neutralizing anti-IFN-I autoAb repertoires at discrete times that selectively targeted various combinations of IFNα, IFNβ, and IFNω. Longitudinal analyses further revealed that emergence of neutralizing anti-IFNα autoAbs correlated with reduced IFN-stimulated gene (ISG) levels, indicating impairment of innate immunity. Patient data review suggested that prior recorded viral infections and autoimmune history influence the likelihood of mounting anti-IFN-I autoAbs. Indeed, systematic measurements in biobanked samples revealed significant enrichment of pre-existing autoreactivity against clinically relevant autoantigens in individuals who later developed anti-IFN-I autoAbs. In this context, we describe lifelong neutralizing anti-IFNα autoAbs (and impaired innate immunity), that manifested in an individual following IFNα therapy, and who was retrospectively found to have had pre-existing autoreactivity to β2-glycoprotein-I before IFNα treatment. Our decades-spanning longitudinal analyses illuminate the development and immune implications of anti-IFN-I autoAbs in an aging population, and support a ‘two-hit’ hypothesis whereby loss of self-tolerance prior to immune-triggering with endogenous or exogenous IFN-I may pose a risk for developing late-onset, lifelong IFN-I functional deficiency.

## INTRODUCTION

Deficiencies in the human type I interferon (IFN-I) system leave individuals susceptible to a range of severe viral diseases typically caused by pathogens to which they lack pre-existing humoral immunity (reviewed in (Duncan et al., 2021; Meyts and Casanova, 2021; Stertz and Hale, 2021)). While life-threatening viral diseases linked directly to genetic defects in the IFN-I system are extremely rare (and mainly manifest themselves in the very young) (Duncan et al., 2021), it has recently become apparent from cross-sectional studies that a functional defect caused by autoantibodies (autoAbs) targeting IFN-I cytokines is not particularly rare in elderly populations (reviewed in (Bastard et al., 2024a; Hale, 2023)). Specifically, the prevalence of autoAbs neutralizing the IFN-I cytokines IFNα and/or IFNω increases sharply with age in otherwise healthy individuals, such that a conservative estimate of prevalence in those >70 years old is about eight times higher (1.4%) than that in younger individuals (0.17%) (Bastard et al., 2021). Furthermore, the presence of neutralizing anti-IFN-I autoAbs in individuals has been associated with an increased susceptibility to severe infections caused by several viral pathogens, including SARS-CoV-2 (Akbil et al., 2022; Bastard et al., 2024b; Bastard et al., 2020; Busnadiego et al., 2022; Chauvineau-Grenier et al., 2022; Credle et al., 2022; Eto et al., 2022; Frasca et al., 2022; Goncalves et al., 2021; Manry et al., 2022; Mathian et al., 2022; Scordio et al., 2022; Solanich et al., 2021; Troya et al., 2021; Wang et al., 2021), MERS-CoV (Alotaibi et al., 2023), influenza A virus (Zhang et al., 2022), West Nile virus (Gervais et al., 2023; Lin et al., 2023), and various herpesviruses (Bayat et al., 2015; Burbelo et al., 2010; Busnadiego et al., 2022; Hetemaki et al., 2021; Mathian et al., 2022; Mogensen et al., 1981; Pozzetto et al., 1984). Given that more than 100 million people worldwide have been estimated to potentially harbor neutralizing anti-IFN-I autoAbs (Bastard et al., 2024a), and are therefore at increased risk of severe infectious disease outcomes, it is critical to understand factors associated with their development and pathogenic mechanisms in order to inform future mitigation strategies.

Multiple distinct host genetic defects converging on disruption of central T-cell tolerance in the thymus have been shown to underlie the development of anti-IFN-I autoAbs in several patient cohorts (reviewed in (Bastard et al., 2024a; Hale, 2023)). The best characterized examples of these defects include mutations in the *AIRE* gene, which encodes an autoimmune regulator that normally ensures negative selection of autoreactive T-cells (Meager et al., 2006; Meyer et al., 2016), and mutations in the *NFKB2, MAP3K14* (NIK) and *RELB* genes, that encode components of the alternative NF-κB pathway and regulate *AIRE* expression (Bodansky et al., 2022; Le Voyer et al., 2023; Ramakrishnan et al., 2018; Sjogren et al., 2022). Patients with these genetic defects exhibit thymic abnormalities and reduced self-tolerance, and typically develop anti-IFN-I autoAbs in early childhood (Le Voyer et al., 2023; Meager et al., 2006), which has been speculated to occur following an infection event triggering ‘immunization’ with endogenous IFN-I (Hale, 2023). However, the role of genetics and other pre-disposing factors in contributing to the increased prevalence of anti-IFN-I autoAbs in elderly populations is little understood, and it still remains to be resolved at the individual level at what point in life these autoAbs develop, their longevity, and their impact on an individual’s innate antiviral defenses. Furthermore, associations between potential ‘immunization’ events, such as infections, and the development of anti-IFN-I autoAbs have yet to be investigated. These gaps in our knowledge mainly result from a lack of available longitudinal samples and lack of systematic clinical histories taken from before anti-IFN-I autoAbs developed in individuals, as well as the limited time that has passed so far for patients recently identified to harbor anti-IFN-I autoAbs, which has prevented long-term follow-up studies.

Here, we sought to leverage the Swiss HIV Cohort Study (SHCS) as a large nationwide, systematic, longitudinal infectious disease cohort to study the development and consequences of anti-IFN-I autoAbs in elderly individuals. The SHCS was founded in 1988, and contains historic, semiannually biobanked plasma and cell samples, as well as clinical data, from more than 21,000 people living with HIV (PLH) over the course of 35 years (Scherrer et al., 2022). With improved success of long-term antiretroviral therapy, life-expectancy in this cohort is now approaching that of the general population in Switzerland (Gueler et al., 2017). Thus, this well-treated cohort is aging, and individuals >50 years of age make up around 60% of the currently enrolled patients, with most having been followed-up for almost half their adult lives. Furthermore, IFN-I therapy was widely used in this cohort for hepatitis C virus (HCV) treatment before directly acting antivirals became available (Baumann et al., 2024). Using this resource, we wanted to track the levels of neutralizing and non-neutralizing anti-IFN-I autoAb repertoires (targeting IFNα, IFNβ, and IFNω) in individuals over decades at high-resolution, thus providing key information on the timing of autoAb induction and their longevity. Furthermore, we aimed to describe the impact of neutralizing anti-IFN-I autoAbs on innate antiviral defenses, and to perform exploratory analyses of clinical record data to investigate whether pre-disposing factors or events influence the likelihood of anti-IFN-I autoAb development.

## RESULTS

### Identification and characterization of anti-IFN-I autoAbs in an aging, longitudinally-sampled, infectious disease patient cohort

To systematically investigate the age-related development of anti-IFN-I autoAbs in the SHCS, we initially selected recent plasma samples taken from a sub-cohort of 1876 well-treated individuals >65 years of age (82% male, median year of birth 1949, IQR 1943-1954; **Supplementary Table 1**). Age and sample availability were the only criteria considered in this selection. We then applied a multiplexed bead-based assay to screen these samples for IgG autoAbs binding to the representative type I IFNs: IFNα2, IFNβ and IFNω. Samples identified to be positive were subsequently tested for neutralization capacity against the respective IFN-I at different doses. We observed that 0.85% of individuals (16/1876) had neutralizing autoAbs against IFNα2, 0.32% of individuals (6/1876) had neutralizing autoAbs against IFNβ, and 0.48% of individuals (9/1876) had neutralizing autoAbs against IFNω (**Fig. 1A**). Overall, 1.17% of the individuals in our >65 cohort (22/1876) had neutralizing autoAbs against at least one IFN-I, a prevalence similar to that noted previously in similarly-aged, otherwise healthy, individuals (Bastard et al., 2021). An additional 0.69% of individuals in our >65 sub-cohort (13/1876) had detectable anti-IFN-I autoAbs that were non-neutralizing (0.11% IFNα2, 0.21% IFNβ and 0.48% IFNω). With regards patient ethnicity, most likely source of HIV-1 infection, and baseline levels of HIV-1 RNA, there were no clear differences between those with detectable levels of anti-IFN-I autoAbs compared to those without (**Supplementary Table 1**). However, males made up 94.3% (33/35) of those identified to have anti-IFN-I autoAbs as compared to 81.9% (1507/1841) of males in the population without anti-IFN-I autoAbs, suggesting a trend towards increased prevalence in males (**Supplementary Table 1**). Notably, one individual (P5) had IgG autoAbs binding and neutralizing all three type I IFNs, while most had binding autoAbs specific for IFNα2 (0.53%, 10/1876), IFNω (0.43%, 8/1876), or both IFNα2 and IFNω (0.37%, 7/1876) (**Figs. 1B & 1C**). Seven individuals (0.37%) only had IgG autoAbs specific to IFNβ, while two individuals (0.11%) had IgG autoAbs against IFNβ and IFNω (**Figs. 1B & 1C**). While most anti-IFN-I autoAb-positive plasmas neutralized the respective IFN-I, it was striking that the majority of plasmas positive for anti-IFNω autoAbs alone appeared to be non-neutralizing at the plasma dilution tested (**Fig. 1C**). Further characterization of each individual’s anti-IFN-I IgG autoAb subclasses revealed a binding antibody response dominated by IgG1 autoAbs in most individuals (**Figs. 1D & 1E**). However, it was interesting to observe that IgG4 was also a commonly identified subclass of anti-IFNα2 and anti-IFNω IgG autoAbs (statistically significant for IFNα2, but not for IFNω), and that IgG4 even dominated in a few individuals with these types of autoAbs. In contrast, it was notable that anti-IFNβ IgG4 did not dominate in any individual (**Fig. 1E**). Anti-IFN-I IgG autoAbs of the IgG3 subclass were also sporadically detected, and represented the main IgG autoAb in some individual patients, while anti-IFN-I IgG2 autoAbs were very rarely detected or of low titer (**Figs. 1D & 1E**). Thus, the anti-IFN-I IgG autoAb landscape in our sub-cohort is mostly comprised of the IgG1 subclass. Overall, our screening data reveal the diversity of anti-IFN-I autoAbs in a sub-cohort of PLH individuals >65 years of age, and indicate that the prevalence of these autoAbs in this sub-cohort (1.17% for neutralizing autoAbs only) is highly comparable to that previously reported in a similarly-aged healthy cohort (Bastard et al., 2021).

**Figure 1.**
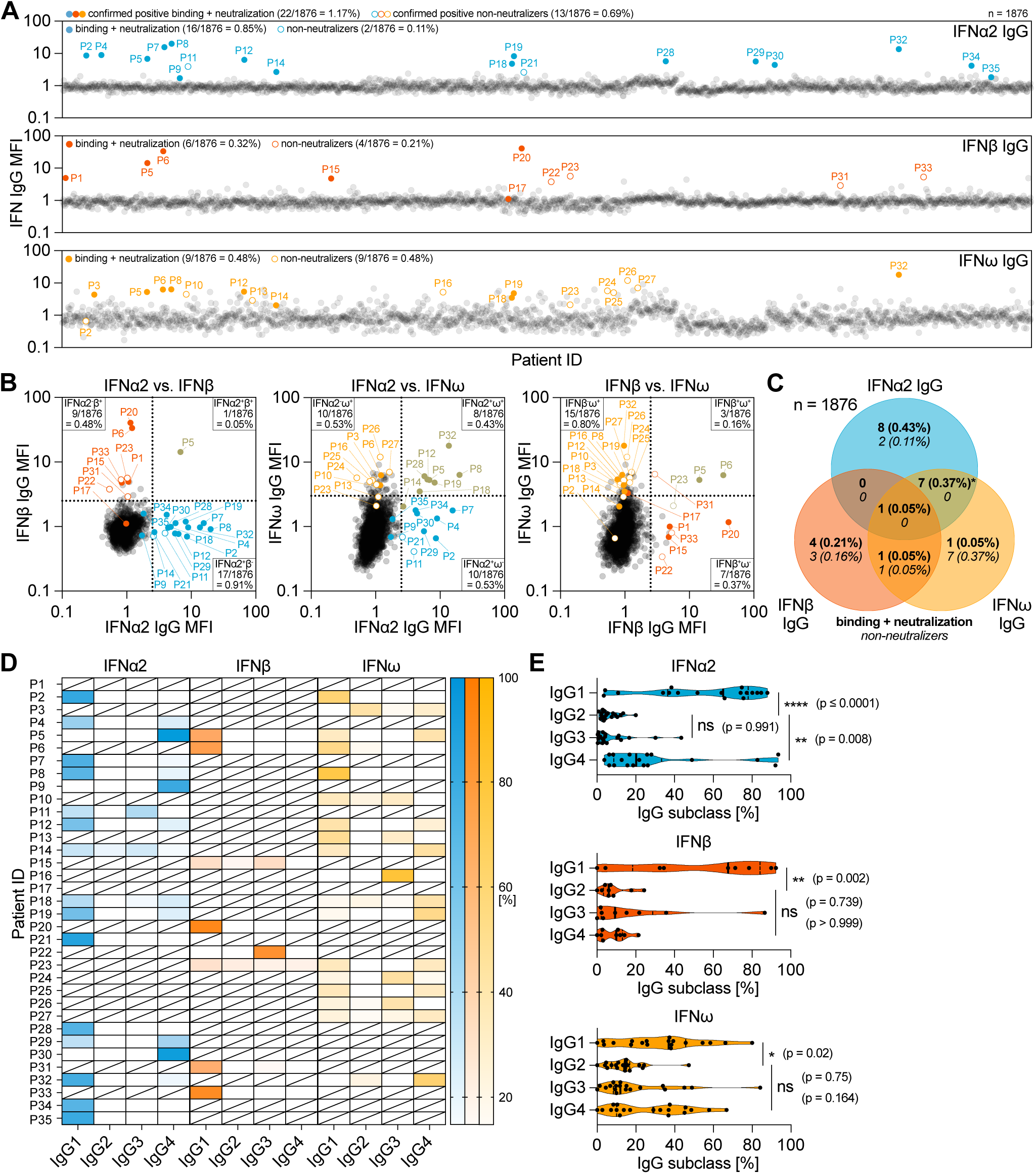
Identification and characterization of anti-IFN-I autoAbs in an aging, longitudinally-sampled, infectious disease patient cohort. **(A-C)** Validated screening results for presence of anti-IFNα2, anti-IFNβ and anti-IFNω IgG in plasma samples derived from 1876 unique patients enrolled in the SHCS and aged >65 years at the time of sampling. Median Fluorescence Intensity (MFI) Fold Change (FC) of IgG values obtained from IFN-I-coated beads relative to the MFI of IgG values obtained from empty beads is shown, normalized to the cohort means for each IFN-I. All individual patient samples are shown (circles), with samples considered positive after subsequent analysis of longitudinal samples colored (see Methods for thresholds). Solid colored circles represent plasma samples that also neutralized the respective IFN-I in subsequent assays. Numbers and percentage of positive patients (neutralizing and non-neutralizing IgG) are indicated for each anti-IFN-I IgG. (B) Pairwise representation of the data shown in (A) comparing the indicated combinations of anti-IFNα2, anti-IFNβ and anti-IFNω IgG found in each patient plasma sample. (C) Venn diagram analysis of the 35 anti-IFN-I autoAb-positive patients highlighting the anti-IFN-I autoAb specificities observed for neutralizing and non-neutralizing IgG. Percentages refer to the entire sub-cohort (1876 patients). The asterisk denotes the inclusion of a single patient found to possess binding and neutralizing anti-IFNα2 IgG, as well as binding and non-neutralizing anti-IFNω IgG. **(D-E)** Plasma samples from anti-IFN-I autoAb-positive patients were analyzed to determine relative levels (%) of each of the four IgG subclasses targeting IFN-Is. (D) Patient-level analysis shown as a heat-map, where white blocks indicate no anti-IFN-I IgG subclass was detected, and slashed blocks indicate IgG subclass was not determined. (E) IgG subclass analysis in all patients for each IFN-I. Statistical analysis was performed using a one-way ANOVA with Tukey’s multiple comparison (single pooled variance). *P* values and significance are indicated (ns = non-significant).

### High-resolution longitudinal analysis of anti-IFN-I autoAb levels reveals their acute age-associated development and persistence over decades

For all 35 anti-IFN-I autoAb positive individuals, as well as for 35 anti-IFN-I autoAb negative individuals (matched for sex and year of birth), we obtained historic biobanked plasma samples from all timepoints available in the SHCS since enrollment. Samples were typically available from two timepoints (approximately 6-monthly) per year, and spanned an average of 20.2 years (range 9-27) for each anti-IFN-I autoAb positive individual. As an example, the anti-IFN-I autoAb positive individual with the most samples available had 52 plasma samples that were taken between the ages of 43 and 70 (i.e. over a 27-year period). For all longitudinal plasma samples we assessed levels of IgG autoAbs binding to each type I IFN, and subsequently tested most samples for IFN-I neutralization capacity, resulting in a temporal overview of anti-IFN-I autoAb binding and neutralization development for each individual (**Fig. 2 and Supplementary. 1**). Despite individual heterogeneity with regard to anti-IFN-I autoAb reactivity, several common features were apparent from analyzing the anti-IFN-I autoAb positive population. Firstly, for 34/35 anti-IFN-I autoAb positive individuals, anti-IFN-I autoAbs were undetectable for many years in the available samples prior to the occurrence of a positive sample, indicating that the autoAbs did not exist in these individuals for most of their lives. This is in stark contrast with patients harboring mutations in the *AIRE* gene or in genes of the alternative NF-κB pathway, where anti-IFN-I autoAbs are detectable very early in childhood (Le Voyer et al., 2023; Meager et al., 2006). Indeed, confirming the age-related onset of anti-IFN-I autoAbs in our study population, the median ages of new-onset autoAb detection were remarkably similar, at 63, 63, and 61.5 years for anti-IFNα2, anti-IFNβ, and anti-IFNω, respectively, with a range of ages spanning years 45-80 (**Fig. 2A**). Secondly, anti-IFN-I autoAbs first developed at a specific, discrete timepoint in each individual, indicating that their induction was likely triggered by a certain acute event. Thirdly, anti-IFN-I autoAbs generally increased in binding titers over time for anti-IFNα2 and anti-IFNω, but not for anti-IFNβ (**Fig. 2B**), which might reflect increases in autoAb abundance or increases in avidity towards the IFNα2 and anti-IFNω antigens. However, it is unclear why this appears to be antigen specific. Finally, with the exception of rare transient ‘blips’ at early timepoints (e.g. P8), it was apparent that the presence of anti-IFN-I autoAbs never resolved, and once developed they persisted continuously in subsequent plasma samples (**Figs. 2C-I and Supplementary. 1**). For example, the longest duration of neutralizing anti-IFN-I autoAbs detected in our study sub-cohort was approximately 15 years (P12 and P32; **Fig. 2D**), and 11 individuals maintained anti-IFN-I autoAbs for at least 10 years each. On the contrary, anti-IFN-I binding autoAbs were never detected at any timepoint in the 35 negative individuals tested longitudinally (see examples in **Fig. 2I and Supplementary. 2**). Overall, our longitudinal data reveal acute induction of anti-IFN-I autoAbs that can occur in some individuals around the age of 60-65 years, which is followed by subsequent lifelong maintenance of these anti-IFN-I autoAbs.

**Figure 2.**
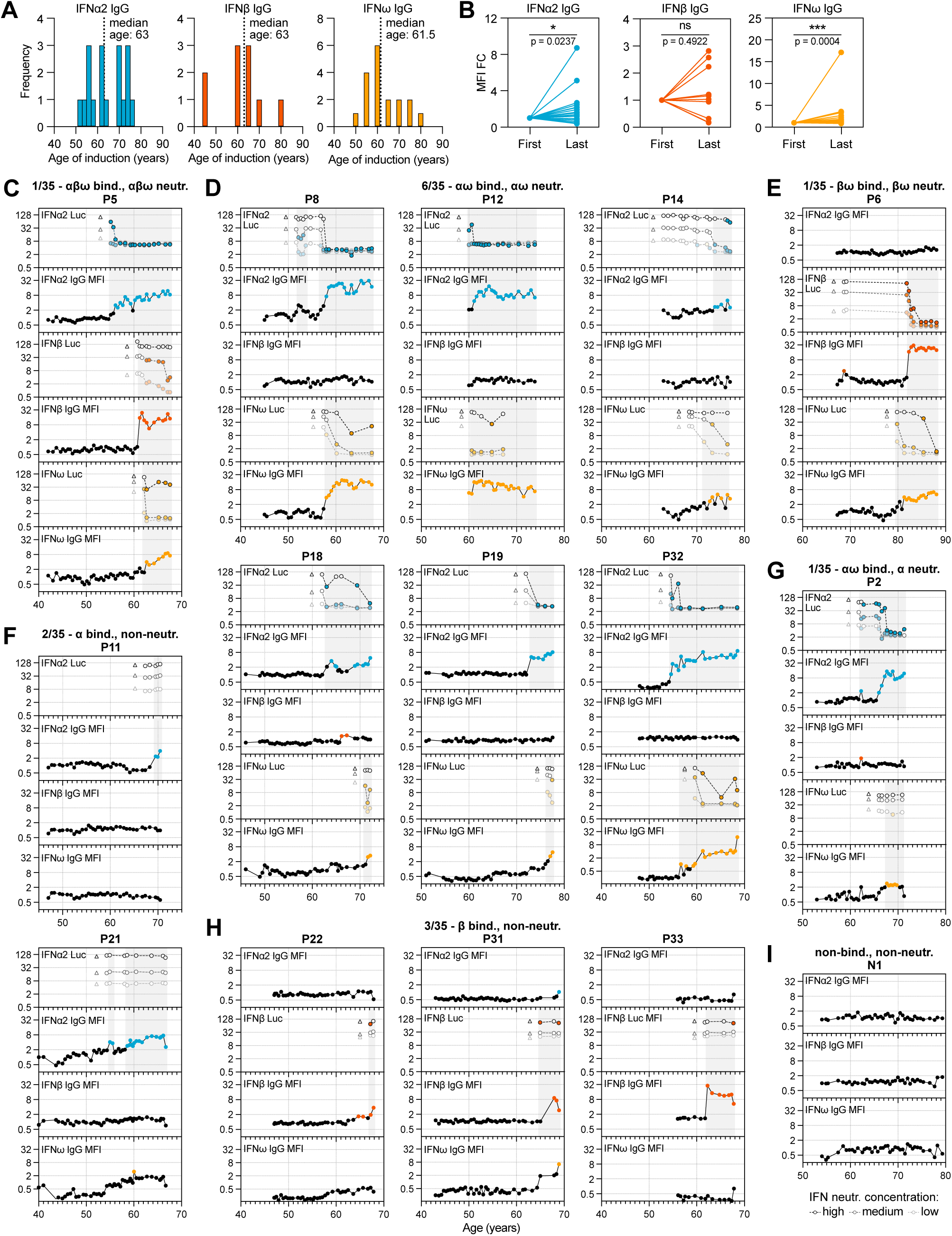
High-resolution longitudinal analysis of anti-IFN-I autoAb development over decades. Semiannually biobanked plasma samples available for all 35 anti-IFN-I autoAb-positive patients (and several negative patients) were analyzed for anti-IFNα2, anti-IFNβ and anti-IFNω IgG levels, as well as for IFNα2, IFNβ or IFNω neutralization capacity. **(A)** Frequency of ages (years) where each anti-IFN-I autoAb was first detected. The median age of first detection (induction) is noted. **(B)** MFI FC values for each anti-IFNα2, anti-IFNβ and anti-IFNω IgG in each patient comparing relative levels between the first timepoint where anti-IFN-I autoAbs were detected and the last available timepoint sampled. Statistical analysis was performed using a Wilcoxon matched-pairs signed rank test. *P* values and significance are indicated (ns = non-significant). **(C-I)** Patient-level representation of anti-IFNα2, anti-IFNβ and anti-IFNω IgG levels (MFI FC), as well as IFNα2, IFNβ or IFNω neutralization (inhibition of IFN-induced Luciferase (Luc) activity) at 3 different doses (see Methods), for all available longitudinal samples plotted as a function of patient age (years). Colored circles represent samples considered positive for either binding IgG or neutralization (see Methods for thresholds). Triangles in neutralization plots represent negative controls. In (C-I), individual patients are grouped according to the types of IFN-I to which they possess binding and neutralizing IgG (indicated at the top of each panel, together with the n/35 patients who have a similar phenotype). The patient in (I) is a negative patient who never developed anti-IFN-I autoAbs. See also Supplementary Figures 1 and 2.

### Distinct individual variation in the timing and specificity of anti-IFN-I autoAb development

We observed various phenotypes relating to anti-IFN-I autoAbs that were particular to certain individuals, or to groups of individuals. For example, it was striking in the single individual with autoAbs binding and neutralizing all three type I IFNs (P5) (**Fig. 2C**) that each anti-IFN-I autoAb developed at different times over the course of seven years, with anti-IFNα2 autoAbs developing after age 55, anti-IFNβ autoAbs developing after age 61, and anti-IFNω autoAbs developing after age 62. This suggests that different acute events might have triggered development of these different neutralizing autoAb reactivities at different times. A similar pattern was observed in individual P32, who developed neutralizing anti-IFNα2 autoAbs after age 55, followed by neutralizing anti-IFNω autoAbs around age 60 (**Fig. 2D**). However, anti-IFNβ autoAbs never developed in P32, highlighting inter-individual variability in the types of IFN-I antigens to which anti-IFN-I autoAbs can be raised. While reactivity to both IFNα2 and IFNω was not uncommon to observe (7/35 individuals; **Figs. 2D** and **2G**) there were many examples where reactivity to either only IFNα2 (10/35 individuals; e.g. **Fig. 2F** and **Supplementary. 1C**) or IFNω (8/35 individuals; **Supplementary** Figs. 1B and 1E) occurred, indicating that specificity of reactivity at the individual level, even between these two closely related type I IFNs, is possible despite the known potential for IFNα/IFNω cross-reactive autoAbs to exist (Meyer et al., 2016). Furthermore, such lack of reactivity to both IFNα2 and IFNω was not simply a consequence of short time periods preventing the broadening of reactivity from one IFN to the other, as some individuals had specific anti-IFNα2 or anti-IFNω autoAbs for 8 years (P4, P21; **Fig. 2F** and **Supplementary. 2C**), 11 years (P7), or even 15 years (P10; **Supplementary. 2E**) without developing autoAbs against the other IFN. In a similar specificity example, individual P6 developed anti-IFNω autoAbs after the age of 80 and then developed anti-IFNβ autoAbs after the age of 82, but never developed anti-IFNα2 autoAbs (**Fig. 2E**). Thus, even within 35 individuals, almost all combinations of anti-IFN-I autoAb reactivities could be observed, which also extended to variations of whether the autoAbs were able to neutralize the action of IFN-I or not. Overall, our longitudinal analyses reveal that neither time of induction, binding specificity, nor neutralization capacity of anti-IFN-I autoAb repertoires can be generalized, but probably reflect distinct host or environmental factors specific to each individual.

### Development of neutralizing anti-IFN**α**2 autoAbs is associated with compromised baseline IFN-stimulated gene levels

Our production of high-resolution longitudinal anti-IFN-I autoAb binding and neutralization data in a highly-studied infectious disease cohort, combined with the availability of biobanked samples and clinical data spanning multiple decades, gave us the possibility to dissect consequences of anti-IFN-I autoAb development in individual patients. Given the heterogeneity of types of anti-IFN-I autoAbs observed in our sub-cohort, we focused studies on individuals who developed neutralizing anti-IFNα2 autoAbs, with the rationale that this was the largest ‘homogeneous’ sub-group (n=16), and that neutralization capacity was most likely to have explainable functional consequences. We began by looking at the consequences of anti-IFN-I autoAb development on well-documented database-recorded outcomes related to HIV-1 viral loads and consequences, incidences of other viral infections (e.g. herpes zoster, HSV-1 and CMV), bacterial and fungal infections, and outcomes such as diabetes or cancers. For each individual, we analyzed these outcomes for their whole lifetime following first detection of neutralizing anti-IFNα2 autoAbs, and compared the results to anti-IFN-I autoAb negative individuals within the SHCS database who were matched for sex, registration date, study center, and year of birth (n=62). We did not observe a significant effect of neutralizing anti-IFNα2 autoAbs on any of the recorded outcomes (**Supplementary Table 2**) which, for HIV-1 at least, might reflect the high effectiveness of long-term antiretroviral therapy. Furthermore, at the cellular level, we did not observe that development of neutralizing anti-IFNα2 autoAbs correlated with any changes to specific blood cell compositions, including leukocytes, platelets, or various lymphocyte sub-populations (**Fig. 3A**). To investigate directly whether the development of neutralizing anti-IFNα2 autoAbs had consequences for IFN-stimulated gene (ISG) expression, we next analyzed baseline ISG levels in multiple frozen biobanked PBMC samples that were available from 13 individuals who developed neutralizing anti-IFNα2 autoAbs, as well as from 13 age-matched negative-control individuals who never developed anti-IFN-I autoAbs. For each autoAb-positive individual, we obtained 2-3 PBMC samples from timepoints before, and 2-3 PBMC samples from timepoints after, autoAb development. In the case of negative-control individuals, we obtained a similar number of samples from age-matched timepoints. Without culturing, total RNA was extracted directly from frozen PBMCs and subjected to RT-qPCR analysis for a panel of 8 well-known ISG mRNAs: *MX1*, *RIGI*, *IRF9*, *RSAD2*, *IFITM3*, *IFIT2*, *IFIT3*, and *IFI44*. As shown in **Fig. 3B**, mRNA levels of all ISGs were significantly reduced in autoAb-positive individuals subsequent to their development of neutralizing anti-IFNα2 autoAbs. In contrast, baseline ISG mRNA levels were not reduced in autoAb-negative individuals at age-matched timepoints, and more than half of these individuals actually exhibited increased levels of several ISGs over time (**Fig. 3C**). Overall, these results build on previous cross-sectional descriptions of a correlation between neutralizing anti-IFNα autoAbs and low ISG levels (Abers et al., 2021; Bastard et al., 2020; Kisand et al., 2008; Koning et al., 2021; Lopez et al., 2021; van der Wijst et al., 2021; Wang et al., 2021), and reveal that age-associated development of neutralizing anti-IFNα2 autoAbs can have clear temporal functional consequences, leading to compromised baseline ISG levels and thus impaired innate antiviral responses.

**Figure 3.**
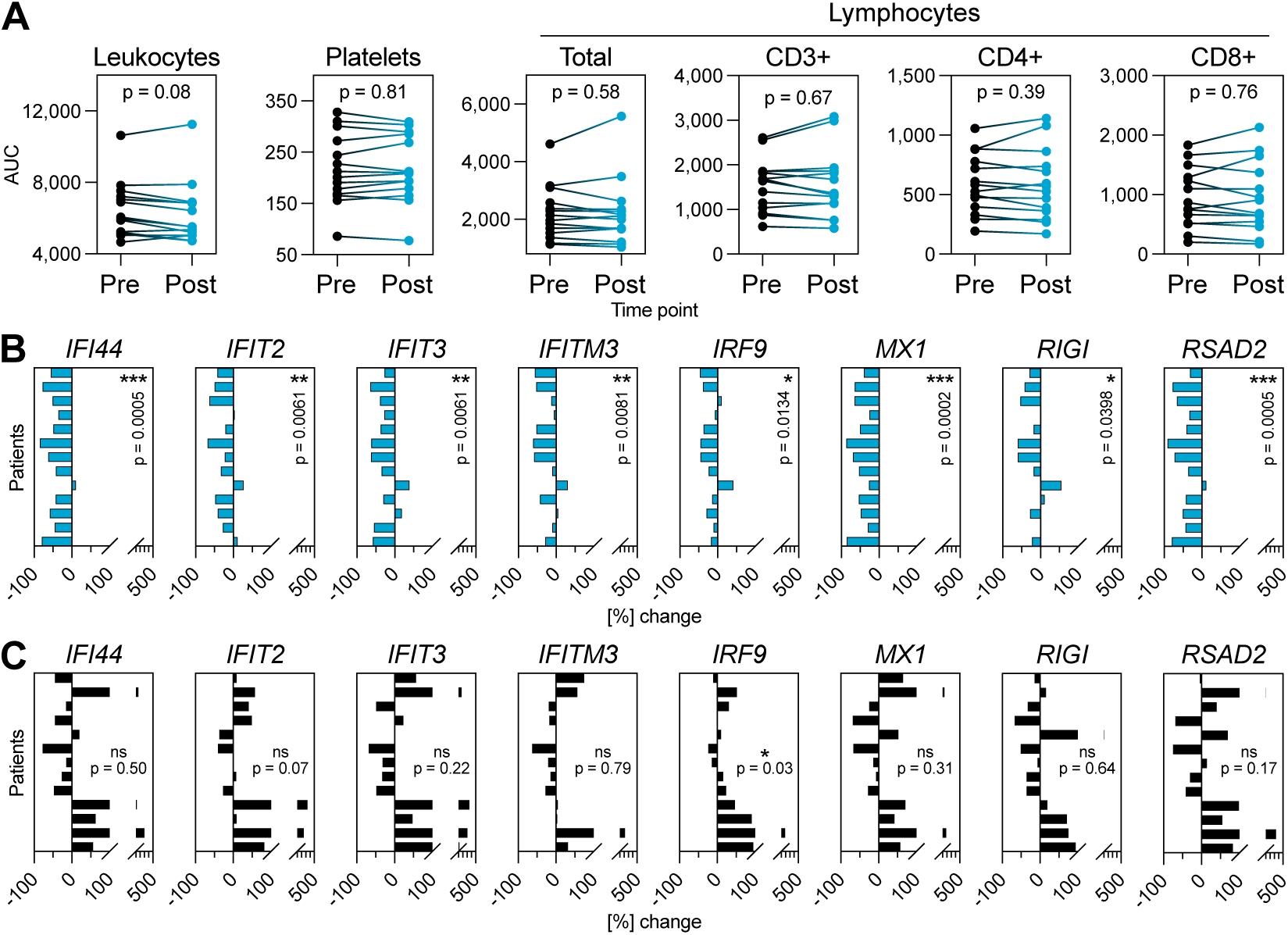
Neutralizing anti-IFNα2 autoAbs are associated with compromised baseline IFN-stimulated gene (ISG) levels. **(A)** Area under the curve (AUC) values for clinically determined cell compositions in whole blood (as indicated) in 16 patients who developed neutralizing anti-IFNα2 autoAbs. Existing clinical cell titers were obtained for each patient from the SHCS, and AUC values were determined from all available data up to 1 year before (Pre) or 1 year after (Post) the timepoint where anti-IFNα2 autoAbs were first detected. Statistical analysis was performed using a paired Wilcoxon signed rank test. *P* values are indicated. **(B-C)** RT-qPCR analysis of the indicated ISGs in PBMCs from 13 patients who developed neutralizing anti-IFNα autoAbs (B) or 13 age-matched control patients who never developed anti-IFN-I autoAbs (C). Data shown for each patient represent mean percentage changes in expression of the indicated ISG relative to the first timepoint (i.e. samples taken before development of anti-IFNα autoAbs for (B), or to the equivalent timepoint for (C)). Statistical significance of changes across the 13 patients was determined based on the original ΔCt values (normalized to *GAPDH*) using a Mann-Whitney U test. *P* values and significance are indicated (ns = non-significant).

### Prior virus infections may influence anti-IFN-I autoAb development

Beyond understanding the functional consequences of anti-IFN-I autoAbs, our longitudinal dataset gave us the unique opportunity to probe the clinical records of each patient and to understand factors potentially influencing the development of anti-IFN-I autoAbs. To do this, we initially analyzed differences between our 35 anti-IFN-I autoAb positive patients and 138 autoAb negative patients (matched for sex, registration date, study center, and year of birth) at all timepoints that preceded first onset of anti-IFN-I autoAbs in positive individuals. Given the matching for registration date and year of birth, the same timepoints could be considered for the negative individuals. Out of 30 parameters investigated in an exploratory study, which were either chosen due to their routine recording in the SHCS database or their infection-relatedness, only prior CMV seropositivity and prior diagnosis of herpes zoster showed significant differences between the two groups (**Supplementary Table 3** and **Figs. 4A-B**). Notably, prior CMV seropositivity had a small negative association with anti-IFN-I autoAb development (p=0.00696; **Fig. 4A**), suggesting a potential protective effect of CMV infection on anti-IFN-I autoAb development. Indeed, CMV seropositivity has previously been reported to also negatively associate with the development of multiple sclerosis (Sundqvist et al., 2014; Waubant et al., 2011), an autoimmune disease where heterogeneous pathogenic autoAbs may play important roles (Höftberger et al., 2022). Thus, it could be that CMV persistence contributes to subtle immunomodulatory changes to the host that reduce certain autoimmune responses in some otherwise predisposed individuals.

**Figure 4.**
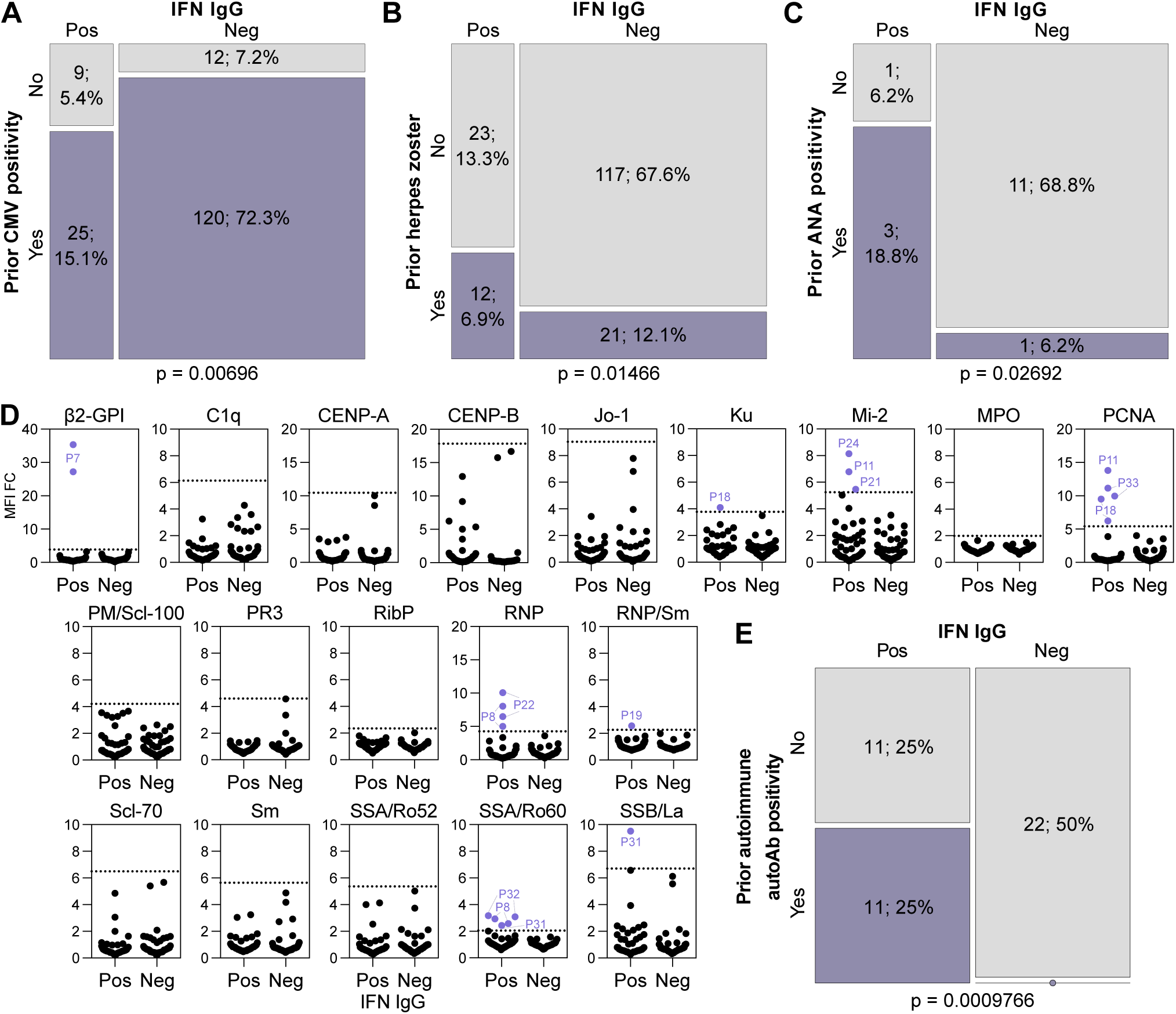
Prior infections and immune factors influence the development of anti-IFN-I autoAbs. **(A-C)** Mosaic plots comparing the SHCS recorded incidence of prior CMV positivity (A), prior herpes zoster diagnosis (B), or prior ANA test positivity (C) between patients who developed anti-IFN-I autoAbs and matched control patients who did not. In (A) and (B), only patients with complete data for the indicated parameter were included (therefore n differs slightly with that in Supplementary Table 3). In (C), only patients who were tested are included. **(D)** Screening results for the presence of 19 different anti-autoantigen IgGs in plasma samples derived from 22 anti-IFN-I autoAb positive (Pos) patients and 22 age-matched negative control (Neg) patients who were confirmed to have never developed anti-IFN-I autoAbs. Two samples per patient were tested, which were the two samples immediately preceding first detection of anti-IFN-I autoAbs (for the positive patients; typically 6 and 12 months before) or age-matched timepoints for the negative patients. MFI FC IgG values obtained from the indicated autoantigen-coated beads are shown relative to the MFI of IgG values obtained from empty beads normalized to the negative patient mean for each autoantigen. All patient samples are shown (circles). Patient plasmas exhibiting normalized MFI values >5 SDs above the mean MFIs obtained from the negative patient samples (dotted lines) were considered positive for the specific anti-autoantigen IgG and are colored and labelled. **(E)** Mosaic plot analysis of the data in (D). For panels (A) and (B) statistical analyses were performed using conditional-logistic regression (taking into account the matched nature of the data) and likelihood-ratio tests, for panel (C) statistical analysis was performed using Fisher’s Exact Test for Count Data (as the routine ANA data were not available for matched pairs of cases and controls), and for panel (E) statistical analysis was performed using Exact McNemar test (as this takes into account both the paired nature of the data and the absence of events in one group). *P* values are indicated.

Our analyses also showed that prior herpes zoster events had a small positive association with anti-IFN-I autoAb development (p=0.01466; **Fig. 4B**). This was particularly surprising, as further temporal analysis revealed that the prior recorded herpes zoster diagnoses did not always immediately precede the onset of anti-IFN-I autoAb development, as might be expected if the severe acute disease itself triggered autoAb production. Specifically, out of the 12 individuals who developed anti-IFN-I autoAbs and had documented prior herpes zoster, only two individuals (P10 and P24) had herpes zoster in the six months prior to first detection of anti-IFN-I autoAbs. Of the remaining 10 individuals, the herpes zoster events were recorded variously between three and 14 years prior to anti-IFN-I autoAb onset. However, it is possible that the recorded herpes zoster events might act as a well-documented indicator of other disparate types of severe infections that may also have occurred in these patients, perhaps closer to the time of anti-IFN-I autoAb development. In this regard, it is notable that a previous study postulated that a range of recurrent severe or chronic infections may be factors in promoting the development of autoAbs, including against IFN-Is, in certain infection-susceptible immunodeficient individuals (Walter et al., 2015). To explore this further on a case-by-case basis in our sub-cohort, we looked for additional recorded infection-related clinical events that occurred up to one year prior to first detection of anti-IFN-I autoAbs. One individual (P4) was reported to have recently suffered from a severe pneumonia with sepsis that led to hospitalization. Respiratory samples taken from P4 at this time were found to be PCR-positive for influenza A virus and bocavirus, the former agent being paradigmatic for inducing significant IFN-I responses (Dunning et al., 2018). Another individual (P7) was treated for HCV infection with ribavirin and pegylated IFNα, but the normal course of treatment was cut short for an unknown reason, and P7 developed anti-IFNα2 autoAbs during this period. Thus, our finding of a small, yet statistically significant, association between prior herpes zoster diagnoses and anti-IFN-I autoAb development, together with additional case-by-case analyses of individual infection-related events, may support the concept that previous infections might influence autoAb induction (Walter et al., 2015). Mechanistically, as discussed below for the IFNα-treated individual (P7), this could be due to events that increase levels of the IFN-I ‘immunogen’ for subsequent autoAb development, which could include severe infections.

### Pre-existing autoimmune reactivity predicts subsequent anti-IFN-I autoAb development

While exposure to virus-induced IFN-I may act as the ‘immunogen’ for triggering anti-IFN-I autoAb production, a compromised ability to tolerate self-antigens, including IFN-Is, is likely an essential additional requirement for autoAb development. For example, defects in genes relating to correct thymus function, key to self-tolerance mechanisms, have been clearly linked to the early lifetime development of anti-IFN-I autoAbs (Bodansky et al., 2022; Le Voyer et al., 2023; Meager et al., 2006; Ramakrishnan et al., 2018). Furthermore, in the elderly, it may be that common age-related thymic decline plays a key role in reducing self-tolerance and promoting autoAb development in some individuals (Liang et al., 2022). Given the availability of longitudinal clinical data and samples for our sub-cohort, we speculated that further analysis may reveal evidence of general loss of self-tolerance that precedes induction of anti-IFN-I autoAbs in certain patients. We therefore initially searched the available clinical records of patients we had analysed for anti-IFN-I autoAbs to identify instances where their autoimmune status had been assessed by ANA (anti-nuclear antigen) test. While the number of patients tested due to clinical need was small (n=4 for those with anti-IFN-I autoAbs, n=12 for matched controls), there was nevertheless a statistically significant association between prior ANA test positivity and subsequent development of anti-IFN-I autoAbs in our sub-cohort (p = 0.02692; **Fig. 4C**). To test the association between loss of self-tolerance (e.g. ANA positivity) and subsequent anti-IFN-I autoAb induction more formally, we next selected 22 out of our 35 anti-IFN-I autoAb positive patients for whom we had at least 5 years of validated anti-IFN-I autoAb negativity prior to anti-IFN-I autoAb development. For each patient, we obtained the two plasma samples taken immediately before the first confirmed detection of anti-IFN-I autoAbs (this was typically 6 months and 1 year prior), and used a multiplexed bead-based assay to screen these samples for IgG autoAbs binding to 19 different human autoantigens associated with autoimmune disease, including those likely detected by the clinical ANA test. In parallel, age-matched samples from 22 (negative) patients who never developed anti-IFN-I autoAbs were also tested. Using standard deviation thresholds based on these negative patient samples, we identified 11 patients with IgG autoAbs targeting diverse ANA-related and non-related autoantigens, and it was striking that all of these 11 patients (100%) were positive individuals who subsequently went on to develop anti-IFN-I autoAbs (**Fig. 4D**). None of the 22 anti-IFN-I autoAb negative patients exhibited autoreactivity to these other autoantigens. There was no apparent specificity to the observed prior autoreactivity in those who went on to develop anti-IFN-I autoAbs, with different patients harboring IgG targeting distinct autoantigens (including β2-GPI, Ku, Mi-2, PCNA, RNP, RNP/Sm, SSA/Ro60, and SSB/La). However, it is interesting to note that anti-SSA and anti-SSB autoAbs are associated with primary Sjögren’s syndrome where frequent development of anti-IFN-I autoAbs has been reported (Burbelo et al., 2019; Gupta et al., 2016). These data amounted to a statistically highly significant association between prior autoimmune autoAb positivity and subsequent development of anti-IFN-I autoAbs (p = 0.0009766; **Fig. 4E**). Overall, the statistically significant association between prior ANA test positivity and subsequent development of anti-IFN-I autoAbs, together with our experimental identification of autoreactivity in 50% of those who went on to produce anti-IFN-I autoAbs, indicate that diminished self-tolerance can precede anti-IFN-I autoAb development. Furthermore, these data suggest that those with certain autoimmune diseases, which might be detectable by routine assays such as ANA screening, are at higher risk of subsequently developing anti-IFN-I autoAbs.

### Development and lifelong persistence of neutralizing anti-IFN**α** autoAbs potentially triggered by therapeutic IFN**α** in an individual with pre-existing autoimmunity

Based on previous hypotheses (Hale, 2023), and the data presented so far in this manuscript, we postulate that anti-IFN-I autoAbs develop following exposure to unusually high levels of IFN-I (possibly driven by severe infection) in the context of diminished self-tolerance. Towards validating this concept, we followed-up on the case of individual P7, who developed neutralizing anti-IFNα autoAbs in their mid-50s, and maintained them lifelong ever since (>12 years) (**Fig. 5A**). P7 was initially diagnosed with HCV in their mid-40s, and was treated with the standard of care at the time, ribavirin and pegylated IFNα, eight years later. Treatment with pegylated IFNα was started only 5 weeks before neutralizing anti-IFNα autoAbs were first detected, and all previous samples tested (>12 years) were negative for anti-IFNα autoAbs (**Fig. 5A**). This observation is highly suggestive that the treatment of P7 with pegylated IFNα may have stimulated the production of long-lasting anti-IFNα autoAbs in a manner similar to that previously reported in a single case (Jorns et al., 2006). However, such occurrences would seem to be the atypical situation, as although production of anti-IFNα autoAbs has been described to occur in response to recombinant IFNα treatment, production appears to be transient and usually self-resolving, at least in the small numbers of individuals (20-40) tested previously (Bell et al., 1994; Jorns et al., 2006; Ronnblom et al., 1992). To formally address this on a larger scale, we took advantage of our access to plasma samples taken from 300 individuals in the SHCS (including P7; **Supplementary Table 1**) who had been treated with pegylated IFNα, and assessed whether they had detectable levels of anti-IFN-I IgG autoAbs. Two samples per patient were tested: the first sample available after IFNα treatment began (typically around 6 months); and the last sample available (typically 10-20 years later). Only 3/300 individuals tested around 6 months after starting IFNα treatment were confirmed to possess anti-IFNα IgG autoAbs in their plasmas (P7, P37, and P38), and only the plasma of P7 neutralized the activity of IFNα (**Fig. 5B**). For P37 and P38, the presence of anti-IFNα autoAbs was transient, as these autoAbs were not readily detected in the last available plasma samples from these patients, or in any other longitudinal samples available (spanning up to 20 years) (**Figs. 5B-D**). Analysis of the last available plasma samples from our IFNα-treated cohort revealed that only 2/300 individuals possessed anti-IFNα autoAbs in their plasmas at this timepoint, and both neutralized the activity of IFNα (P7 and P36) (**Fig. 5B**). Notably, longitudinal analysis of biobanked plasma samples from P36 (spanning around 15 years) revealed that his induction of anti-IFNα IgG autoAbs occurred about 10 years after IFNα treatment stopped, suggesting that the IFNα treatment was not the acute trigger for anti-IFNα autoAbs in this individual. We also (re)-tested all available longitudinal plasma samples from 5 negative individuals treated with IFNα in case we had missed important sample timepoints. However, despite some low-level transient anti-IFNα IgG autoAb positive ‘blips’, all these IFNα-treated individuals remained negative for long-lasting neutralizing anti-IFNα autoAbs (**Fig. 5E**). Thus, we conclude that development of long-lasting neutralizing anti-IFNα autoAbs in response to IFNα treatment (as observed for P7) is an atypical, rare event.

**Figure 5.**
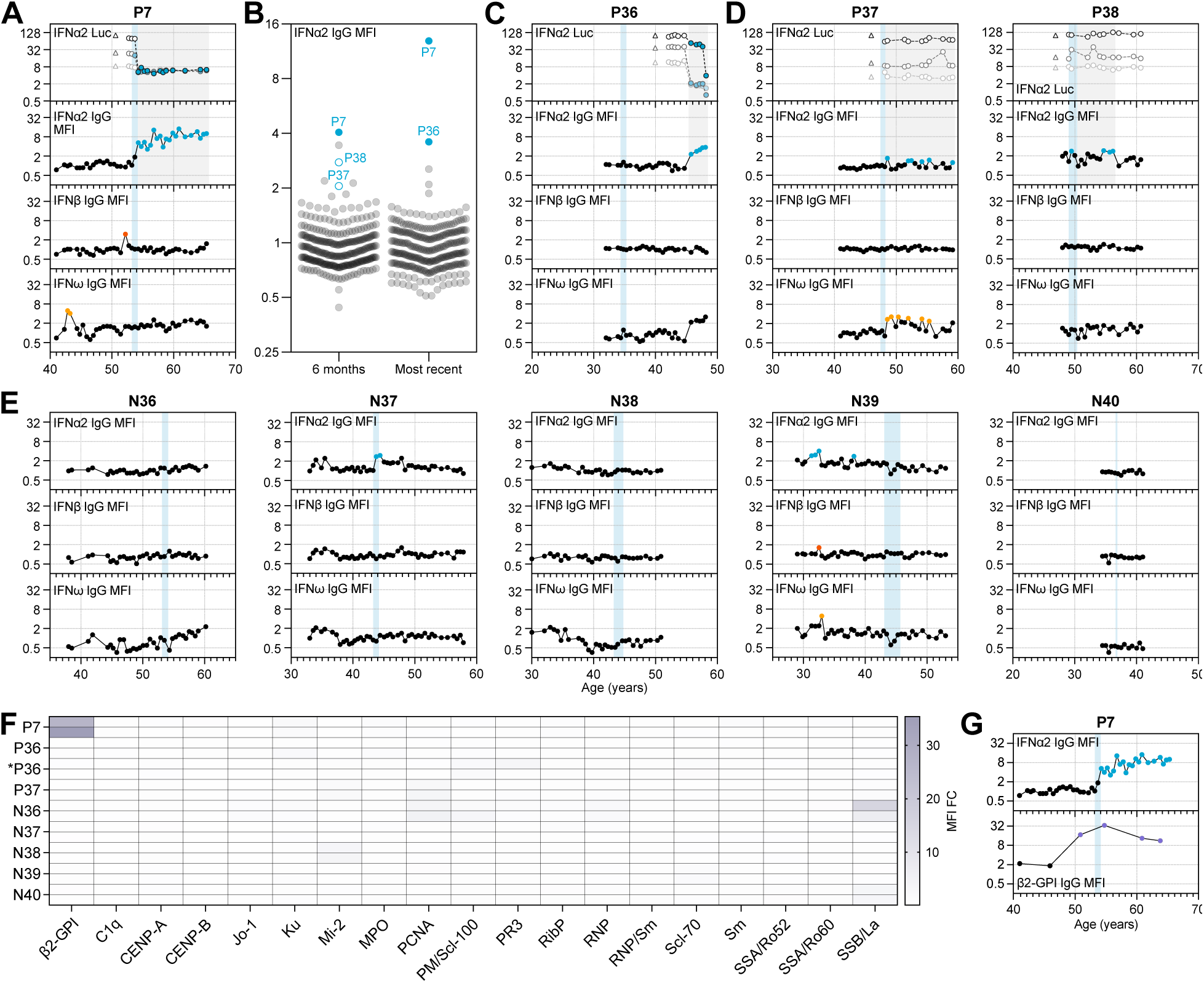
Therapeutic IFNα likely triggered development and lifelong persistence of neutralizing anti-IFNα autoAbs in an individual with pre-existing autoimmunity. **(A)** Representation of anti-IFNα2, anti-IFNβ and anti-IFNω IgG levels (MFI FC), as well as IFNα2 neutralization (inhibition of IFN-induced Luciferase (Luc) activity) at 3 different doses (see Methods), for all available longitudinal samples from patient P7 (who was treated therapeutically with IFNα2) plotted as a function of patient age (years). **(B)** Validated screening results for the presence of anti-IFNα2 IgG in plasma samples derived from 300 unique patients enrolled in the SHCS who were treated with IFNα2. Two samples per patient were assayed: the first sample available after IFNα treatment (typically around 6 months), and the last sample available (most recent: typically 10-20 years later). MFI FC values obtained from IFNα2-coated beads relative to the MFI of values obtained from empty beads is shown, normalized to the cohort mean. All individual patient samples are shown (circles), with samples considered positive after subsequent analysis of longitudinal samples colored (see Methods for thresholds). Solid colored circles represent plasma samples that also neutralized IFNα when assayed. Positive patient samples are labeled. **(C-E)** Representation of data similar to (A), but for all available longitudinal samples from the indicated patients who had also been treated therapeutically with IFNα2. **(F)** Heatmap representation of screening results for the presence of 19 different anti-autoantigen IgGs in plasma samples derived from 8 patients who had been treated therapeutically with IFNα2. Two samples per patient were tested, which were the two samples immediately preceding the start of IFNα2 treatment (typically 6 and 12 months before), as well as immediately preceding first detection of anti-IFN-I autoAbs (for newly-identified patient *P36). MFI FC IgG values obtained from the indicated autoantigen-coated beads are shown relative to the MFI of IgG values obtained from empty beads normalized to the means of controls shown in Fig. 4. Patient plasmas exhibiting normalized MFI values >5 SDs above the mean MFIs obtained from the controls shown in Fig. 4 were considered positive for the specific anti-autoantigen IgG and are colored. Note that the samples for P7 are the same as shown in Fig. 4 as the start of IFNα2 treatment coincided with first detection of anti-IFN-I autoAbs. **(G)** Representation of anti-IFNα2 (from panel A) and anti-β2-GPI IgG levels (MFI FC), in selected longitudinal samples from patient P7 plotted as a function of patient age (years). In panels (A-E) and (G), colored circles represent samples considered positive for either binding IgG or neutralization (see Methods for thresholds). Triangles in neutralization plots represent negative controls. Blue shading indicates the period of time when each patient underwent IFNα2 treatment.

Towards understanding why P7 (but none of the other IFNα-treated individuals) developed a long-lasting neutralizing anti-IFNα IgG autoAb response during treatment, we obtained the two plasma samples taken immediately prior to IFNα treatment (typically 6 months and 1 year prior) for seven IFNα-treated individuals in addition to P7. We then used a multiplexed bead-based assay to screen these samples for IgG autoAbs binding to a panel of different autoimmune disease-associated autoantigens as a means to identify those with potential breakdown of self-tolerance at these timepoints. Notably, P7 had strong unambiguous evidence for autoimmune reactivity (anti-β2-GPI IgG autoAbs) in his plasma samples prior to developing anti-IFNα autoAbs, while autoreactivity was generally lacking in samples taken from all other patients who did not develop anti-IFNα autoAbs following IFNα treatment (except N36 who had weak reactivity to SSB/La) (**Fig. 5F**). Furthermore, longitudinal analysis revealed that P7 possessed anti-β2-GPI IgG autoAbs at least 3 years prior to receiving IFNα treatment and developing long-lasting neutralizing anti-IFNα IgG (**Fig. 5G**). Taken together with our previous observation of a statistically significant association between prior autoreactivity positivity and subsequent anti-IFN-I autoAb development, the results of this case-study suggest that a pre-existing breakdown of self-tolerance in P7 was likely to have been decisive in him mounting a long-lasting anti-IFNα IgG response when treated with pegylated IFNα.

## DISCUSSION

Herein, we systematically investigated the age- and IFN-I-treatment -related development, long-term dynamics, and longevity of anti-IFN-I autoAbs in well-treated individuals enrolled over several decades in the Swiss HIV Cohort Study (SHCS). The unique aspect of our work is the longitudinal analysis of historic biobanked patient samples and clinical records that span the decades before and after each patient developed anti-IFN-I autoAbs. Together with retrospective serological and cellular immunoassays, this allowed us to dissect factors associated with anti-IFN-I autoAb induction and their consequences. Our observation that around 1.9% of individuals developed anti-IFN-I autoAbs (1.17% for neutralizing autoAbs only), with a median onset age of ∼63 years (range 45-80), aligns exceedingly well with findings from a recent cross-sectional study that noted a sharp increase in neutralizing anti-IFN-I autoAb prevalence (to 1.4%) in otherwise healthy individuals over the age of 70 years, as compared with younger individuals (Bastard et al., 2021). This suggests that the underlying HIV-positive status of individuals within the sub-cohort we studied, the majority of whom were well-treated with long-term antiretroviral therapy, likely has only a limited impact (if any) on anti-IFN-I autoAb development. Our results should therefore also be broadly applicable to the otherwise healthy general aging population, and our data now provide a new high-resolution long-term perspective on this form of IFN system deficiency and its contribution to severe viral disease.

The induction of anti-IFN-I autoAbs in some individuals around ages 60-65 is enigmatic. Such late-onset development is suggestive that, at least in this population, germline host genetic variants compromising thymic function may play only a limited contribution as compared with the situation in other younger patient populations described to harbor mutations in *AIRE* or alternative NF-κB pathway genes (Le Voyer et al., 2023; Meager et al., 2006). Nevertheless, our finding that there is a significant temporal association between prior autoimmune reactivity (e.g. via a positive clinical ANA test) and the subsequent development of anti-IFN-I autoAbs is still indicative that these patients may indeed have lost their full self-tolerance capacity. While we cannot rule out an impact of the HIV-1 infection on patient autoimmune status (Roszkiewicz and Smolewska, 2016; Zandman-Goddard and Shoenfeld, 2002), we note that the observed anti-IFN-I autoAb prevalence in our sub-cohort is very similar to that in otherwise healthy individuals (Bastard et al., 2021). Given the genetic link between central T-cell tolerance in the thymus and anti-IFN-I autoAb development (Le Voyer et al., 2023; Meager et al., 2006), the most likely explanation in our aged patients is therefore that the natural process of thymic involution has led to reduced thymus activity and thus an overall increased likelihood of general development of autoimmunity (Liang et al., 2022), including against IFN-Is. Further studies will have to address this hypothesis directly, as well as understand why only a subset of aged individuals go on to develop such autoimmune reactions. In this regard, the availability of longitudinal biobanked plasma and PBMC samples, together with high-resolution tracking of anti-IFN-I autoAb induction (such as presented here), could prove invaluable to retrospectively assess thymus-related immunosenescence in specific aging individuals by assaying historic thymic output (e.g. by quantifying T-cell receptor excision circles, TRECs; (Mitchell et al., 2010)) and correlating this directly with the subsequent likelihood of anti-IFN-I autoAb induction. Given the apparent severe consequences of anti-IFN-I autoAbs for infection susceptibility (reviewed in (Bastard et al., 2024a; Hale, 2023)), in the future there may be a clear clinical role for performing diagnostic autoimmune (e.g. ANA test and others) or TREC quantifications to help predict an individual’s risk of developing this form of IFN system deficiency.

While aging and prior autoimmune reactivity appear to pre-dispose an individual to develop anti-IFN-I autoAbs, the ‘two-hit’ hypothesis suggests that (auto)immunization with IFN-I is a subsequently required trigger in this process (Hale, 2023). Our finding of a statistical association (albeit weak) between prior recorded virus infections such as reactivated herpes zoster, which activates a clear endogenous IFN-I response in patients (Vandoren et al., 2024), and development of anti-IFN-I autoAbs provides some support to the idea that autoimmunization may occur. Similar support comes from our anecdotal finding that anti-IFN-I autoAbs developed in a patient immediately following hospitalization with severe pneumonia possibly caused by influenza A virus, again a scenario where endogenous IFN-I and inflammatory responses will be activated (Dunning et al., 2018). We also cannot rule out that a subset of our patients had specific sub-clinical undiagnosed systemic autoimmune diseases, such as primary Sjögren’s syndrome or systemic lupus erythematosus, which provided an environment where both loss of tolerance and atypically increased levels of IFN-I triggered anti-IFN-I autoAb development (Burbelo et al., 2019; Gupta et al., 2016; Mathian et al., 2022; Trutschel et al., 2022). However, additional evidence supporting the ‘two-hit’ hypothesis comes from our analysis of 300 individuals who were treated (or ‘immunized’) with pegylated IFNα, where we identified that a single individual with unambiguous evidence for loss of self-tolerance prior to therapy was the only individual to generate long-lasting neutralizing anti-IFNα IgG in response to the exogenous IFNα. Overall, this could suggest that some previous acute infections in tolerance-compromised individuals might lead to development of anti-IFN-I autoAbs, and could thus have long-term consequences for enhancing susceptibility to future severe infections. This concept may also apply to some individuals who are still treated therapeutically with IFN-Is (EASL, 2023), and given the ‘two-hit’ hypothesis it may be worth considering future implementation of diagnostic autoimmune pre-screening and risk-benefit evaluations to assess the likely short- and long-term outcomes associated with IFN-I treatment specifically in those with poor self-tolerance.

In contrast to previous cross-sectional studies (Abers et al., 2021; Bastard et al., 2020; Kisand et al., 2008; Koning et al., 2021; Lopez et al., 2021; van der Wijst et al., 2021; Wang et al., 2021), our access to high-resolution longitudinal samples gave us the unique opportunity to study the within-individual consequences of neutralizing anti-IFN-I autoAb development. Compared to samples taken from before anti-IFN-I autoAbs were detectable, we observed a clear temporal correlation between induction of neutralizing anti-IFNα2 autoAbs and a reduction of baseline ISG levels in patient PBMCs. It is logical to assume that the appearance and maintenance of these neutralizing autoAbs led to this functional impairment of innate antiviral immunity, thus contributing to an increased susceptibility to severe viral infections as previously reported (reviewed in (Bastard et al., 2024a; Hale, 2023)). The consequences of harboring other neutralizing anti-IFN-I autoAbs (e.g. against IFNβ or IFNω) could not be readily discerned in our sub-cohort due to the relatively low numbers of positive individuals identified, although a recent report suggests that neutralization of IFNα may have greater pathogenic consequences than neutralization of IFNω (Bastard et al., 2024b). Nevertheless, a key finding from our work was the observation that following induction, neutralizing anti-IFN-I autoAbs can be effectively maintained in the circulation lifelong (over at least 15 years in one individual), meaning that the consequences of anti-IFN-I development could result in extremely long-lasting IFN-I functional deficiency and increased susceptibility to infection. Transient induction of anti-IFN-I autoAbs has been described in some infection scenarios (Chang et al., 2021; Steels et al., 2022), as well as in this study, and could potentially have important physiological regulatory functions such as dampening immunopathologies (Babcock et al., 2024). However, a distinction should be made with the class of long-lasting, lifelong neutralizing anti-IFN-I autoAbs that we mainly describe here, and which functionally impair innate antiviral immunity.

Our study has some limitations, including the retrospective nature and potential biases inherent in long-term cohort studies. For example, as the SHCS was not specifically established to study clinical aspects of autoAbs, autoimmune results (e.g. ANA tests) were only available for some patients that were performed based on clinical need. However, we were able to address this specific deficiency through systematic measurements made retrospectively from biobanked samples. Furthermore, not all (viral) infections can be recorded in an observational cohort study, and the low prevalence of neutralizing anti-IFN-I autoAbs limits the power to assess their impact on subsequent infections. While we believe that the results from our work should also be broadly applicable to otherwise healthy general populations, we also acknowledge that all of our study patients are people living with HIV, and we thus cannot rule out potential confounding factors related to the infection, unknown opportunistic co-infections, or the effects of antiretroviral therapy that could limit the generalizability of our study. In this regard, a major limitation to studying the functional consequences of anti-IFN-I autoAbs was actually that we could not effectively assess their impact on HIV-1 replication due to the majority of patients being on long-term antiretroviral therapy and therefore exhibiting virological suppression at the time of anti-IFN-I autoAb detection. Our initial sampling strategy (>65 years of age) may also have biased our assessment of anti-IFN-I autoAb lifelong persistence, and we may not yet have a true appreciation of transient anti-IFN-I autoAb induction. While future investigations with larger and more diverse cohorts are therefore clearly essential, we also highlight the unique resource that the SHCS represents for studies such as this given the rarity of decade-spanning longitudinally-sampled aging cohorts focused on infectious diseases with the numbers of patients required to reveal uncommon anti-IFN-I autoAbs.

In summary, our study provides a comprehensive longitudinal exploration of the age-related development and specificities of anti-IFN-I autoAbs. Our work identifies prior infection and autoimmune factors that influence the likelihood of anti-IFN-I autoAb induction, and thereby supports the hypothesis that at least ‘two-hits’ probably underlie this process. These findings have implications for diagnosing, perhaps through routine screening of ‘at-risk’ individuals with known autoimmune conditions, those who may be pre-disposed to develop anti-IFN-I autoAbs and the lifelong IFN-I functional deficiency that results. Broader knowledge in this area should contribute to the development of targeted strategies to mitigate severe viral disease susceptibility.

## MATERIALS AND METHODS

### Patient samples, data and ethics

The 1876 plasma samples from patients >65 years at the time of initial testing (**Supplementary Table 1**), 300 plasma samples from patients treated with pegylated IFNα (**Supplementary Table 1**), and all retrospective follow-up longitudinal samples (including plasma and/or PBMCs) analyzed in this study were derived from samples stored in the biobanks of the Swiss HIV Cohort study (SHCS). The SHCS is a prospective, nationwide, longitudinal, noninterventional, observational, clinic-based cohort with semiannual visits and blood collections, enrolling all PLH in Switzerland since 1988 (Scherrer et al., 2022). The SHCS maintains comprehensive, longitudinal, anonymous data collections for all participants, including extensive clinical and demographic data. The data are collected by the five Swiss university hospitals, two cantonal hospitals, 15 affiliated hospitals and 36 private physicians (listed in http://www.shcs.ch/180-health-care-providers). Detailed information on the study is available at http://www.shcs.ch. The SHCS has been approved by the ethics committees of all participating institutions (Kantonale Ethikkommission Bern, Ethikkommission des Kantons St. Gallen, Comite Departemental d’Ethique des Specialites Medicales et de Medicine Communataire et de Premier Recours, Kantonale Ethikkommission Zürich, Repubblica et Cantone Ticino–Comitato Ethico Cantonale, Commission Cantonale d’Étique de la Recherche sur l’Être Humain, Ethikkommission beider Basel), and written informed consent has been obtained from all participants. The personnel who conducted the work with patient samples had no information on patient demographics at the time of analysis, and all data have subsequently been analyzed anonymously.

### Analysis of Autoreactive IgG in Plasma Samples

A previously described high-throughput multiplexed bead-based assay was implemented to assay patient plasma samples for anti-IFN-I IgG (Achleitner et al., 2024; Busnadiego et al., 2022; Liechti et al., 2018). Briefly, magnetic beads (MagPlex-C Microspheres, Luminex) were coupled to recombinant carrier-free human IFN-Is (IFNα2, Novus Biologicals; IFNβ, PeproTech; or IFNω, Novus Biologicals) at a concentration of 10µg protein per million beads, or left empty. Bead coating efficiency and bead specificity were assessed using mouse monoclonal antibodies as described previously (Busnadiego et al., 2022). For serological testing, obtained patient plasma samples were heat-inactivated at 56°C for 1h before being diluted 1:50 in PBS supplemented with 1% BSA (PBS/BSA) and incubated in 96-well plates with 1:1:1:1 mixtures of IFNα2:IFNβ:IFNω:empty beads (2×10^3^ beads per region per well; 1:100 final concentration of plasma). On all experimental plates, a human polyclonal anti-IFNα2b antiserum (BEI resources) was used as a positive control, and an in-house healthy donor pool of human plasmas was used both as a negative control and for batch normalization. Following incubation of the bead:plasma mixes for 1h at room temperature, beads were washed twice with PBS/BSA before phycoerythrin (PE)-labeled secondary antibodies were added separately in PBS/BSA (pan-IgG, 1:500 dilution, Southern Biotech; IgG1, 1:500 dilution, Southern Biotech; IgG2, 1:100 dilution, Southern Biotech; IgG3, 1:100 dilution, Southern Biotech; IgG4, 1:100 dilution, Southern Biotech). After a further 1h incubation at room temperature, bead mixtures were washed twice in PBS/BSA and samples were analyzed on a FlexMap 3D instrument (Luminex). A minimum of 50 beads per antigen were acquired. For each plasma sample, Median Fluorescence Intensity (MFI) values from the IFN-I-coated beads were obtained and made relative to the corresponding MFI value obtained from the empty beads (fold over empty, FOE). Values were then normalized between plates and cohorts using results from the in-house healthy donor pool. In initial screenings, patient plasmas exhibiting normalized FOE values >2 MFI were considered positive for the specific anti-IFN-I IgG. Available longitudinal plasma samples were then obtained for re-testing of all positive patients and selected negative patients. When assaying longitudinal samples, patient plasmas exhibiting normalized FOE values >10 standard deviations (SDs) above the mean MFIs of the first 5 available samples from each individual patient were considered positive for the specific anti-IFN-I IgG.

To assay patient plasma samples for IgG targeting various human autoantigens associated with autoimmune diseases, a customized 19 autoantigen-containing MILLIPLEX® Human Autoimmune Autoantibody Panel (Merck Millipore) was used. The assay was performed according to the manufacturer’s protocol using a 1:300 final dilution of each heat-inactivated patient plasma, and samples were analyzed on a FlexMap 3D instrument (Luminex). For each plasma sample, FOE MFI values for IgG targeting each autoantigen were calculated as described above. Within the dataset for each autoantigen, FOE values were then normalized to the mean MFI values obtained from the anti-IFN-I autoAb-negative patient samples. Patient plasmas exhibiting normalized FOE values >5 SDs above the mean MFIs obtained from the anti-IFN-I autoAb-negative patient samples were considered positive for the specific anti-autoantigen IgG.

### Analysis of IFN-I Neutralization in Plasma Samples

IFN-I neutralization was assessed as described previously using a 293T transient-transfection based dual-luciferase reporter assay in 96-well plates that is based around co-transfection of a plasmid containing the firefly luciferase (FF-Luc) gene under control of the IFN-inducible mouse *Mx1* promoter (pGL3-Mx1P-FFluc) (kindly provided by Georg Kochs) with a plasmid constitutively expressing the *Renilla* luciferase (Ren-Luc) gene (pRL-TK-Renilla) (Busnadiego et al., 2022). Twenty-four hours post-transfection of both plasmids, heat-inactivated patient plasma samples (or multiple negative controls) were diluted 1:50 in DMEM supplemented with 10% fetal calf serum (FCS), 100 U/mL penicillin and 100 mg/mL streptomycin, and incubated for 1h at room temperature with 10, 1 or 0.2 ng/mL of IFNα2 or IFNω, or with 1, 0.2 or 0.04 ng/mL of IFNβ, prior to their addition to the transfected cells. After 24h, cells were lysed for 15 min at room temperature and FF-Luc and Ren-Luc activity levels were determined using the Dual-Luciferase Reporter Assay System (Promega) and a PerkinElmer EnVision plate reader (EV2104) according to the manufacturers’ instructions. FF-Luc values were normalized to Ren-Luc values and then to the median luminescence intensity of control wells that had not been stimulated with any IFN-I. Patient plasmas that reduced IFN-I-stimulated FF-Luc/Ren-Luc values by >2 SDs below the mean values of the negative control samples were considered to be neutralizing.

### Analysis of ISG Expression in PBMC Samples

Thirteen patients who developed persistent neutralizing anti-IFNα autoAbs (positive patients) were selected, together with 13 age-matched control patients who were confirmed longitudinally to have never developed anti-IFN-I autoAbs (negative patients). For each positive patient, we obtained biobanked frozen PBMC samples corresponding to 2-3 annual donations given before development of anti-IFNα autoAbs and 2-3 annual donations given after development of autoAbs. For each negative patient, we obtained similar biobanked frozen PBMC samples corresponding to equivalent timepoints. Total RNA from freshly thawed PBMCs was extracted using the ReliaPrep™ RNA Cell Miniprep System (Promega), and mRNA was subsequently reverse transcribed into cDNA with an oligo(dT) primer using SuperScript III Reverse Transcriptase (Thermo Fisher Scientific) according to the manufacturer’s instructions. Real-time PCR was then performed with PowerTrack™ SYBR Green Master Mix (Thermo Fisher Scientific) using gene-specific forward and reverse primers (sequences available upon request) in a 7300 Real-Time PCR System (Applied Biosystems). Relative gene expression was calculated with the ΔΔCt method. Data were normalized to *GAPDH* levels, averaged between the 2-3 annual donations per patient and per timepoint, and were expressed as percent change relative to the first timepoint (i.e. before development of anti-IFNα autoAbs for the positive patients, or to the equivalent timepoint for the age-matched negative patients). Statistical significance was determined based on the ΔCt values (normalized to *GAPDH*) using a Mann-Whitney U test.

### Statistical Analyses

Patient clinical and demographic data derived from the SHCS relating to the content of Supplementary Tables 1-3, blood cell compositions, prior CMV positivity, prior herpes zoster diagnoses, and prior ANA test positivity were examined using the Wilcoxon rank-sum test (for continuous variables), or Fisher’s exact test (for categorical variables). In addition, conditional logistic regression models were performed to take into account the effect of matching cases and controls. Analyses were performed using R version 4.2.2., with statistical tests noted in the appropriate figure legends. All other statistical analyses presented were performed in GraphPad Prism 9 using the statistical tests noted in the appropriate figure legends.

## Data Availability

All data produced in the present work are contained in the manuscript

## ACKNOWLEDGMENTS

We thank the patients participating in the SHCS and their physicians and study nurses for patient care. The members of the SHCS are Abela IA, Aebi-Popp K, Anagnostopoulos A, Battegay M, Bernasconi E, Braun DL, Bucher HC, Calmy A, Cavassini M, Ciuffi A, Dollenmaier G, Egger M, Elzi L, Fehr J, Fellay J, Furrer H, Fux CA, Günthard HF (President of the SHCS), Hachfeld A, Haerry D (Deputy of ‘Positive Council’), Hasse B, Hirsch HH, Hoffmann M, Hösli I, Huber M, Jackson-Perry D (patient representative), Kahlert CR (Chairman of the Mother & Child Substudy), Kaiser L, Keiser O, Klimkait T, Kouyos RD, Kovari H, Kusejko K (Head of Data Centre), Labhardt N, Leuzinger K, Martinez de Tejada B, Marzolini C, Metzner KJ, Müller N, Nemeth J, Nicca D, Notter J, Paioni P, Pantaleo G, Perreau M, Rauch A (Chairman of the Scientific Board), Salazar-Vizcaya L, Schmid P, Speck R, Stöckle M (Chairman of the Clinical and Laboratory Committee), Tarr P, Trkola A, Wandeler G, Weisser M, and Yerly S. We are grateful to Nadine Rist, Stefan Schmutz, and Kevin Steiner (University of Zurich, Switzerland) for technical support and sample management. We are also grateful to Alexandra Calmy (University of Geneva, Switzerland) and to Matthias Cavassini (University of Lausanne, Switzerland) for their support as representatives of SHCS centers. Financial support for this study was provided by the Swiss National Science Foundation (SNF; grants 31003A_182464 and 310030_214957 to BGH), the Novartis Foundation for Medical-Biological Research (grant 23A069 to BGH), and the Promedica Foundation (to IAA). The study was also co-financed within the framework of the Swiss HIV Cohort Study (SHCS), supported by the SNF (33CS30_201369 to HFG), by the small nested SHCS project 898 (to BGH and IAA). The funders had no role in study design, data collection and analysis, decision to publish, or preparation of the manuscript. The authors declare no competing financial interests.

