## Supplementary figures and images for "Longitudinal Analysis Over Decades Reveals the Development and Immune Implications of Type I Interferon Autoantibodies in an Aging Population"

### Supplemental Figure 1

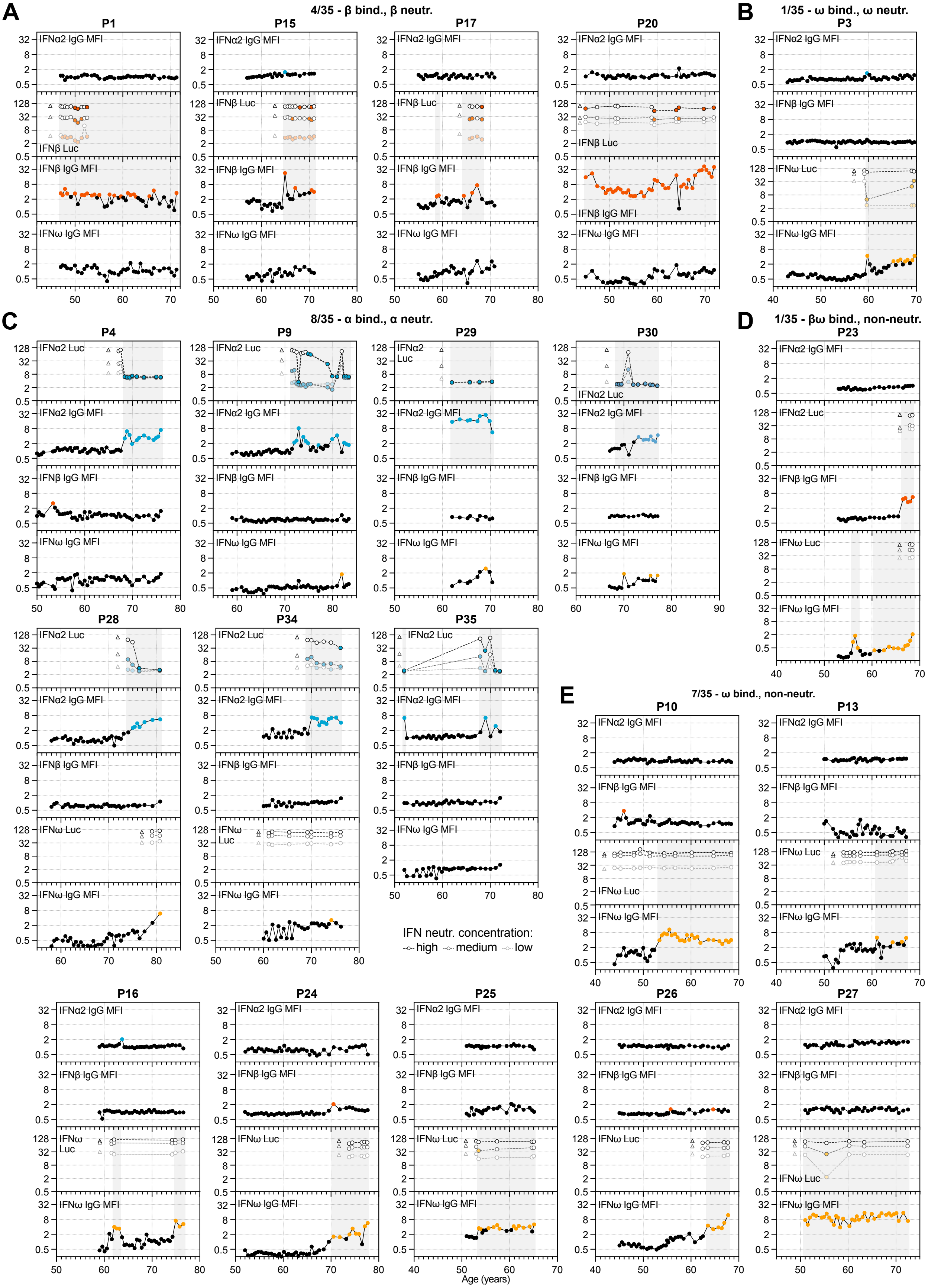

### Supplemental Figure 2

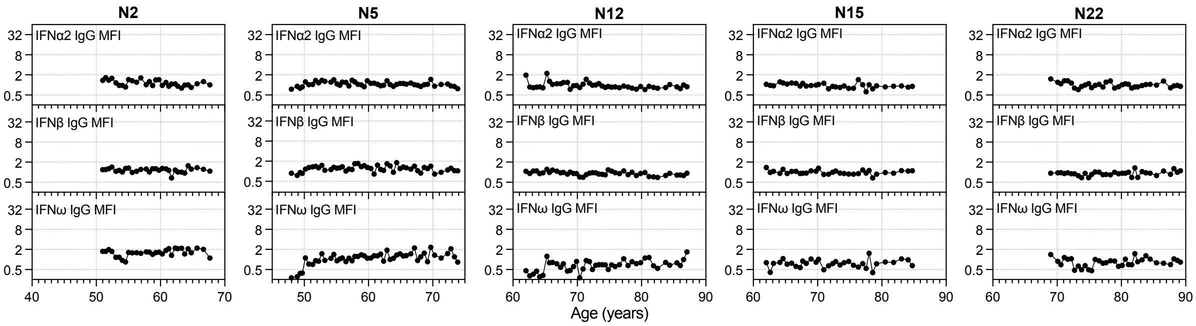
