## Supplemental Tables for "Longitudinal Analysis Over Decades Reveals the Development and Immune Implications of Type I Interferon Autoantibodies in an Aging Population"

### **SUPPLEMENTARY TABLES**

**Supplementary Table 1. Baseline patient characteristics of the study sub-cohorts.**

| **Characteristic** | **Patients >65 yrs with**  **anti-IFN-I autoAbs**  **(n = 35)** | **Patients >65 yrs without**  **anti-IFN-I autoAbs**  **(n = 1841)** | **Patients treated with pegylated IFNα**  **(n = 300)** |
| --- | --- | --- | --- |
|  | **n (%) or median (interquartile range)** | | |
| Year of birth | 1950 (1945-1953) | 1949 (1943-1954) | 1965 (1960-1970) |
| Female | 2 (5.7) | 334 (18.1) | 54 (18.1) |
| Ethnicity (white) | 31 (88.6) | 1704 (92.6) | 281 (94.0) |
| Ethnicity (black) | 3 (8.6) | 84 (4.6) | 5 (1.7) |
| Ethnicity (hispanic) | 1 (2.9) | 16 (0.9) | 3 (1.0) |
| Ethnicity (asian) | 0 (0.0) | 28 (1.5) | 8 (2.7) |
| Ethnicity (other/unknown) | 0 (0.0) | 9 (0.5) | 2 (0.7) |
| Risk group^#^ (HET) | 8 (22.9) | 778 (42.3) | 19 (6.3) |
| Risk group^#^ (IDU) | 3 (8.6) | 73 (4.0) | 164 (54.7) |
| Risk group^#^ (MSM) | 22 (62.9) | 875 (47.5) | 105 (35.0) |
| Risk group^#^ (other/unknown) | 2 (5.7) | 115 (6.2) | 12 (4.0) |
| CD4 (baseline, cells/mm^3^) | 303 (156-525) | 295 (130-487) | 384 (210-572) |
| HIV-1 RNA (baseline, log_10_) | 4.77 (4.07-5.12) | 4.69 (3.81-5.28) | 4.24 (3.47-5.09) |

Abbreviations: IFN-I = type I interferon; autoAbs = autoantibodies; HET = heterosexual; IDU = intravenous drug use; MSM = men who have sex with men.

^#^Risk group refers to most likely source of HIV-1 infection.

**Supplementary Table 2. Impact of neutralizing anti-IFNα autoAbs on recorded outcomes.**

| **Outcome** | **Patients with**  **neutralizing anti-IFNα autoAbs**  **(n = 16)** | **Patients without**  **neutralizing anti-IFNα autoAbs**  **(n = 62)** | ***P* value** |
| --- | --- | --- | --- |
|  | **n (%) or median (interquartile range)** | |  |
| CD4 count (cells/mm^3^) | 612 (400-819) | 548 (395-663) | 0.319^#^ |
| CD8 count (cells/mm^3^) | 530 (331-1079) | 693 (459-1047) | 0.319^#^ |
| Aspergillosis | 0 (0.0) | 1 (1.6) | 1* |
| Bacterial pneumonia | 3 (18.8) | 7 (11.3) | 0.707* |
| Candidiasis (esophageal) | 1 (6.3) | 1 (1.6) | 0.874* |
| Candidiasis (oral) | 0 (0.0) | 2 (3.2) | 1* |
| Diabetes | 1 (6.3) | 6 (9.7) | 1* |
| Encephalopathy (HIV-related) | 0 (0.0) | 2 (3.2) | 1* |
| Herpes simplex (mucocutaneous) | 1 (6.3) | 0 (0.0) | 0.462* |
| Herpes zoster | 0 (0.0) | 5 (8.1) | 0.547* |
| HIV-1 (log_10_ RNA) | 0.00 (0.00-0.00) | 0.00 (0.00-0.00) | 0.949^#^ |
| Mycobacterium avium (disseminated) | 0 (0.0) | 1 (1.6) | 1* |
| Neoplasms | 2 (12.5) | 18 (29.0) | 0.303* |
| Non-Hodgkin’s lymphoma | 0 (0.0) | 2 (3.2) | 1* |
| Pneumocystis pneumonia | 0 (0.0) | 2 (3.2) | 1* |

Abbreviations: IFN-I = type I interferon; autoAbs = autoantibodies.

^#^Wilcoxon rank-sum test

*Fisher’s exact test

**Supplementary Table 3. Impact of prior exposures on development of anti-IFN-I autoAbs.**

| **Prior exposure** | **Patients who developed anti-IFN-I autoAbs**  **(n = 35)** | **Patients who did not develop anti-IFN-I autoAbs**  **(n = 138)** | ***P* value** |
| --- | --- | --- | --- |
|  | **n (%) or median (interquartile range)** | |  |
| Bacterial pneumonia | 0 (0.0) | 7 (5.1) | 0.379* |
| Candidiasis (esophageal) | 3 (8.6) | 7 (5.1) | 0.699* |
| Candidiasis (oral) | 9 (25.7) | 34 (24.6) | 1* |
| Candidiasis (vulvovaginal) | 1 (2.9) | 1 (0.7) | 0.866* |
| Cryptococcosis (disseminated) | 0 (0.0) | 1 (0.7) | 1* |
| Cryptosporidiosis (diarrhea > 1 mo) | 0 (0.0) | 1 (0.7) | 1* |
| **Cytomegalovirus (IgG positivity)^$^** | **25 (71.4)** | **120 (87.0)** | **0.015*** |
| Cytomegalovirus (retinitis) | 0 (0.0) | 2 (1.4) | 1* |
| Diabetes | 1 (2.9) | 16 (11.6) | 0.218* |
| Encephalopathy (HIV-related) | 1 (2.9) | 2 (1.4) | 1* |
| Hepatitis B virus (IgG positivity) | 25 (71.4) | 79 (57.2) | 0.194* |
| Hepatitis C virus (IgG positivity) | 2 (5.7) | 9 (6.5) | 1* |
| Herpes simplex (mucocutaneous) | 1 (2.9) | 0 (0.0) | 0.457* |
| **Herpes zoster^$^** | **12 (34.3)** | **21 (15.2)** | **0.020*** |
| HIV-1 (log_10_ RNA) | 0.00 (0.00-1.64) | 0.00 (0.00-0.00) | 0.199^#^ |
| Kaposi’s sarcoma | 1 (2.9) | 10 (7.2) | 0.574* |
| Microsporidiosis | 0 (0.0) | 1 (0.7) | 1* |
| Mycobacterium avium (disseminated) | 0 (0.0) | 1 (0.7) | 1* |
| Myelopathy (HIV-related) | 0 (0.0) | 1 (0.7) | 1* |
| Neoplasms | 5 (14.3) | 10 (7.2) | 0.324* |
| Neuropathy (peripheral, HIV-related) | 2 (5.7) | 1 (0.7) | 0.195* |
| Non-Hodgkin’s lymphoma | 3 (8.6) | 2 (1.4) | 0.093* |
| Oral hairy leukoplakia | 8 (22.9) | 25 (18.1) | 0.692* |
| Pneumocystis pneumonia | 3 (8.6) | 12 (8.7) | 1* |
| Smoking (baseline, ever) | 15 (42.9) | 84 (60.9) | 0.067* |
| Syphilis (screening test) | 8 (22.9) | 24 (17.4) | 0.674* |
| Toxoplasmosis (screening test) | 18 (51.4) | 80 (58.0) | 0.445* |
| Thrombocytopenia (HIV-related) | 3 (8.6) | 6 (4.3) | 0.563* |
| Tuberculosis (latent) | 2 (5.7) | 5 (3.6) | 0.990* |
| Tuberculosis (pulmonary) | 2 (5.7) | 3 (2.2) | 0.581* |

Abbreviations: IFN-I = type I interferon; autoAbs = autoantibodies.

^#^Wilcoxon rank-sum test

*Fisher’s exact test

^$^For percentage calculations shown here, the indicated total n was used. In the Fig. 4 percentage calculations, n differs slightly as only patients with complete data for the indicated parameter were included
